# Pretreatment Lesion Burden and Cerebrovascular Spatial Context Define Distinct Early Response Trajectories of Brain Metastases After Stereotactic Radiosurgery

**DOI:** 10.64898/2026.11.26.26369087

**Authors:** Yiding Han, Daniel Allan Hamstra, Abdallah Sherif Radwan Mohamed, Ahmed Wagih Mohamed Saber Moawad, Baozhou Sun, Zaid Ali Siddiqui

## Abstract

**Background:** Early imaging changes after stereotactic radiosurgery (SRS) for brain metastases are heterogeneous and can be difficult to interpret clinically. We hypothesized that pretreatment lesion burden and the surrounding cerebrovascular spatial context contribute differently to early post-SRS response.

**Methods:** Individual brain metastases were longitudinally evaluated in two independent clinical cohorts comprising total 116 patients with 513 lesions. Pretreatment lesion volume was measured from three-dimensional segmentations, and early response was categorized as complete response (CR), partial response (PR), stable disease (SD), or progressive disease (PD) based on volumetric change at the first eligible post-SRS MRI within 120 days. A distance-weighted peritumoral vascular field was calculated based on a cerebroarterial probability atlas registered to pretreatment MRI. Associations between lesion volume, vascular field, and early response were assessed using lesion-level longitudinal analyses, including volume-matched PD–PR comparisons and joint volume–vascular response mapping.

**Results:** CR was strongly associated with smaller pretreatment lesion volume, while addition of vascular-field or territorial anatomy information provided little further discrimination. Among 51 volume-matched PD–PR pairs, vascular differences were strongest within the immediate 0–5 mm peritumoral region. In lesions with PD, greater pretreatment vascular field was associated with progressively greater volumetric enlargement (*P* = 0.0106); no corresponding association was observed for the magnitude of PR (*P* = 0.813). Pretreatment lesion volume and vascular context jointly defined partially distinct PR-enriched and PD-enriched states, with a sensitivity of 0.77 (95% CI: 0.65-0.87) compared with 0.64 (95% CI: 0.51-0.75) for the best volume-only threshold, with *P* = 0.04.

**Conclusions:** Pretreatment lesion burden and cerebrovascular spatial context characterize different components of early post-SRS response. Favorable response was predominantly associated with lesion burden and was relatively independent of vascular context, whereas the magnitude of early enlargement increased with the local peritumoral vascular field. These findings support a lesion-level tumor–host framework for understanding post-SRS response heterogeneity and motivate longitudinal studies linking pretreatment vascular context to clinically consequential progression and salvage intervention.

## 1. Introduction

Stereotactic radiosurgery provides effective local therapy for brain metastases, yet interpretation of post-treatment imaging remains a major clinical challenge. An enlarging or increasingly enhancing lesion on surveillance magnetic resonance imaging does not necessarily represent viable tumor progression, as treatment-related changes—including pseudo-progression and radiation necrosis—can closely mimic recurrent disease. Distinguishing these entities is clinically consequential: misclassification may lead to unnecessary invasive intervention or repeat irradiation, inappropriate modification of effective systemic therapy, or, conversely, delayed treatment of true local progression. Conventional response assessment remains largely dependent on serial changes in lesion size and enhancement, which alone cannot reliably distinguish treatment-related enlargement from persistent tumor growth. Understanding why individual brain metastases follow different post-treatment volumetric trajectories may therefore provide biological insight into one of the most difficult problems encountered during surveillance after local therapy.

The cerebral vascular environment is particularly relevant to this post-treatment uncertainty because vascular processes are involved in both tumor persistence and radiation-related tissue injury. Radiation can disrupt endothelial integrity and the blood–brain barrier, alter vascular permeability, and promote edema and inflammatory tissue responses. These mechanisms can produce overlapping radiographic manifestations despite representing fundamentally different biological processes. Accordingly, the spatial relationship between metastasis and the surrounding cerebral vasculature may influence not only its capacity for continued growth after treatment, but also the magnitude and evolution of treatment-related imaging change. Characterizing this vascular context before treatment may therefore provide a complementary perspective on the subsequent trajectory of an individual brain metastasis.

When an enlarging lesion becomes apparent after stereotactic radiosurgery, serial conventional MRI and advanced techniques such as perfusion MRI, diffusion imaging, magnetic resonance spectroscopy, and positron-emission tomography may help distinguish viable tumor from treatment-related change, although no single imaging approach provides definitive classification from pretreatment stage in routine clinical practice.[1, 2] Quantitative MRI and radiomic approaches have also been investigated in relation to treatment outcome [3]; however, many such studies focus on longer-term endpoints such as local control, recurrence, or survival rather than the early lesion trajectory that creates immediate diagnostic uncertainty[4, 5]. Their clinical translation is further limited by sensitivity to image acquisition, preprocessing, segmentation, and cohort-specific characteristics, particularly for the small metastases commonly treated with SRS[6-8]. More fundamentally, these approaches predominantly interrogate the imaging phenotype of the tumor itself and are generally designed to classify outcome rather than to identify the biological or anatomical context underlying early divergence in lesion behavior. Brain metastases are nonrandomly distributed with respect to cerebral anatomy and perfusion, suggesting that their spatial environment contains biologically relevant information; however, whether the pretreatment relationship between an individual metastasis and the cerebral vascular architecture is associated with its early post-SRS evolution remains poorly understood.[9]

We therefore hypothesized that the pretreatment vascular spatial context of an individual brain metastasis is associated with its early longitudinal response after stereotactic radiosurgery. To our knowledge, this is the first study to investigate early post-SRS lesion evolution through the pretreatment spatial relationship between individual brain metastases and the cerebral vascular architecture. We developed an atlas-based, distance-weighted vascular field to quantify this host environment and integrated it with lesion-level longitudinal MRI across independent clinical cohorts. We examined whether vascular context was associated with distinct early response trajectories—including complete response (CR), partial response (PR), stable disease (SD), and progressive disease (PD)—and whether this relationship was distinct from the established effect of pretreatment lesion volume and from categorical territorial anatomy. By shifting the focus from the imaging phenotype of the tumor alone to the pretreatment spatial relationship between tumor and host environment, this study establishes an anatomically interpretable framework for investigating early post-SRS response heterogeneity, particularly during the clinically challenging period when lesion enlargement may reflect either treatment-related change including pseudo-progression (PsP) or persistent tumor progression (TP).

## 2. Methods

### 2.1 Study design and patient cohorts

This retrospective longitudinal study evaluated individual brain metastases from two independent clinical cohorts treated with stereotactic radiosurgery (SRS). For each patient, the pretreatment contrast-enhanced T1-weighted MRI acquired before SRS was designated as the baseline examination *t*_0_ , and the first eligible post-treatment MRI obtained within 120 days after SRS was designated as the early post-SRS examination *t*_1_. The individual metastasis served as the primary unit of longitudinal analysis. The cohorts are further characterized as follows:

1. UCSF BMSR cohort. The University of California, San Francisco Brain Metastasis Stereotactic Radiosurgery (UCSF BMSR) dataset [10] included 306 patients, of whom 95 had both pretreatment and eligible early post-SRS MRI examinations for longitudinal analysis. Tumor contours were manually delineated by expert clinicians as part of routine SRS treatment planning. Early post-SRS lesion contours were additionally reviewed and validated by two attending physicians.
2. MOLAB cohort. The Mathematical Oncology Laboratory (MOLAB) brain metastasis dataset [11] included 75 patients, of whom 31 had both pretreatment and eligible early post-SRS MRI examinations. Tumor contours were manually delineated by expert clinicians as part of routine SRS treatment planning. Early post-SRS lesion contours were additionally reviewed and validated by three attending physicians.

This study was conducted under an institutional review board (IRB)–approved research protocol. All data were obtained from previously published, de-identified datasets, and no additional clinical or demographic information was collected for the present study.

### 2.2 Longitudinal lesion matching and early response assessment

Individual brain metastases were longitudinally matched between the pretreatment *t*_0_ and early post-SRS *t*_1_ examinations based on spatial correspondence on contrast-enhanced T1-weighted MRI and lesion segmentations (segmentation was performed by a previously described pipeline). Lesion volume was calculated from the three-dimensional segmentation at each time point. Pretreatment and early post-treatment lesion volumes were denoted *V*_0_ and *V*_1_, respectively, and relative volumetric change was calculated as

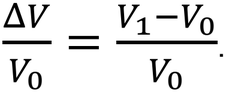

Early treatment response was categorized using volumetric response terminology adapted from the Response Assessment in Neuro-Oncology Brain Metastases (RANO-BM) framework[12]. Lesions that were no longer radiographically identifiable at *t*_1_ were classified as complete response (CR) – these were verified by a human observer. Among lesions remaining measurable at *t*_1_, partial response (PR) was defined as a decrease in lesion volume of at least 30% (Δ*V*/*V*_0_ ≤ −0.30), progressive disease (PD) as an increase in lesion volume of at least 20% (Δ*V*/*V*_0_ ≥ 0.20), and stable disease (SD) as a volumetric change between these thresholds. These definitions are analogous but not identical to RANO which utilizes maximal cross-sectional diameter measurements rather than volume and were chosen to increase discriminatory power.

### 2.3 Cerebral vascular atlas and territorial anatomy mapping

Two complementary population-based cerebrovascular atlases were used to characterize the pretreatment vascular environment of each brain metastasis: a continuous arterial probability map representing the expected spatial distribution of cerebral arteries, and an territorial anatomy atlas defining the major vascular territories of the brain.

The cerebro-arterial probability atlas developed by Mouches and Forkert [13] was generated from TOF-MRA and T1-weighted MRI from 544 healthy subjects acquired across multiple centers and scanners. Cerebral arteries were segmented from individual MRA examinations, nonlinearly normalized to MNI space, and combined to generate a voxel-wise map representing the population probability of arterial occurrence.

A complementary territorial anatomy atlas developed by Liu et al. [14] was constructed from a large clinical cohort of 1,298 patients and partitioned the brain into hierarchical vascular territories, including bilateral anterior cerebral artery (ACA), middle cerebral artery (MCA), posterior cerebral artery (PCA), and vertebrobasilar territories, with finer sub-territorial labels for regional arterial supply.

For each study patient, both atlases were transformed into the pretreatment contrast-enhanced T1-weighted MRI space using affine spatial registration. The continuous cerebroarterial probability atlas was resampled using linear interpolation, whereas the categorical territorial anatomy atlas was resampled using nearest-neighbor interpolation to preserve discrete anatomical labels. Registration results were visually reviewed for anatomical concordance by an attending physician with CNS expertise before lesion-level analysis. A representative example of the registered pretreatment MRI, cerebroarterial probability map, and combined territorial anatomy/vascular representation is shown in Figure 1.

**Figure 1.**
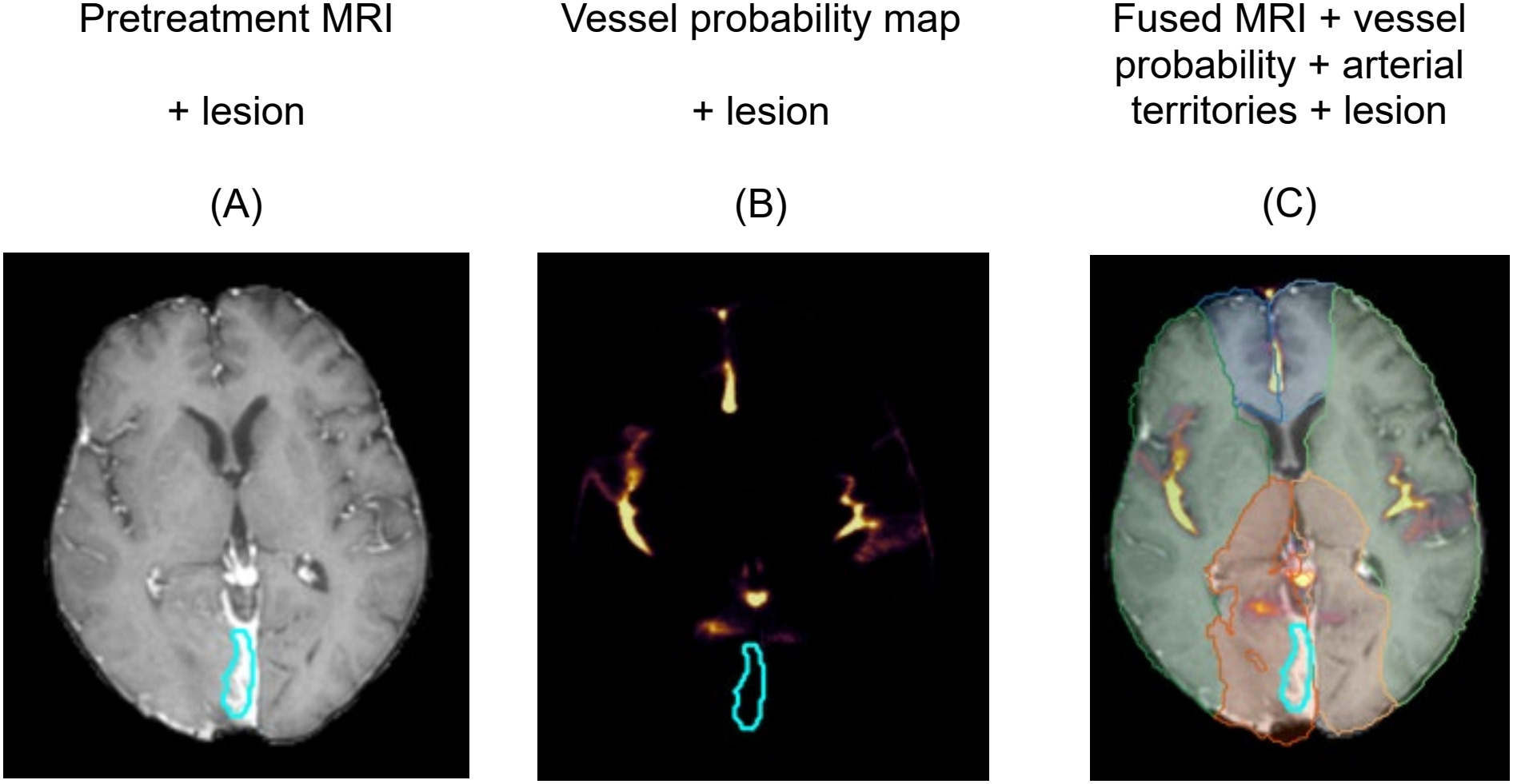
Atlas-based characterization of the pretreatment cerebrovascular environment of an individual brain metastasis. Representative axial images after registration to the pretreatment contrast-enhanced T1-weighted MRI. (A) Pretreatment MRI with the brain metastasis contour shown in cyan. (B) Registered cerebroarterial probability atlas, displayed as a continuous vessel-probability map, with the lesion contour overlaid. (C) Combined visualization of the pretreatment MRI, cerebroarterial probability map, and territorial anatomy atlas, illustrating the lesion’s spatial relationship to both local vascular probability and broader arterial anatomy. Vessel probability is displayed as a percentage according to the color scale.

### 2.4 Quantification of the peritumoral vascular field

To characterize the spatial scale of the peritumoral vascular environment, concentric non-overlapping shells were generated outside the pretreatment lesion surface at 0–5 mm, 5–10 mm, 10–20 mm, and 20–30 mm. Voxels belonging to the index lesion or to another segmented metastasis were excluded. The registered cerebroarterial probability atlas was normalized to a 0–1 scale, such that each voxel represented the population probability of arterial occurrence.

For each lesion, the distance-weighted “vascular field” was calculated by integrating vascular probability over the peritumoral shell:

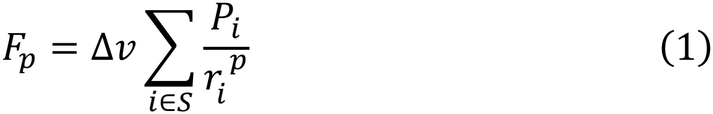

where *P*_*i*_ is the normalized arterial probability at voxel *i* within shell *S, r*_*i*_ is the Euclidean distance from the lesion center of mass to that voxel, Δ*v* is the voxel volume, and *p* determines the degree of spatial weighting. Inverse-distance kernels with *p*=1,2,3 were evaluated across the different peritumoral shells. A minimum distance corresponding to one-half of the smallest voxel dimension was imposed to avoid singular behavior at very short distances.

### 2.5 Statistical analysis

#### 2.5.1 Volume-matched PD–PR comparison

To reduce confounding by pretreatment lesion size, an analysis of lesions matched on volume within +/-20% was performed. A total of 51 matched PD–PR lesion pairs were identified. Within these matched pairs, vascular field differences were evaluated across the predefined distance kernels and spatial shells using the two-sided Wilcoxon signed-rank test. This analysis was used to determine the spatial scale at which vascular context most strongly differentiated PD from PR and to identify the primary vascular-field definition used in subsequent analyses.

#### 2.5.2 Vascular field and volumetric response

Among lesions remaining measurable at early follow-up, the association between the primary vascular-field metric and continuous relative volume change, Δ*V*/*V*_0_ , was evaluated separately for PD and PR lesions. Analyses of vascular-field associations were restricted to lesions with a non-zero pretreatment peritumoral vascular field. Linear regression was used to estimate the direction and magnitude of association, with slopes, 95% confidence intervals, and two-sided *P* -values reported. Binned medians and interquartile ranges were used for descriptive visualization.

The joint distribution of pretreatment lesion volume and vascular field was examined across response categories. A monotonic empirical boundary was constructed using piecewise cubic Hermite interpolation in log-transformed volume space, with boundary parameters optimized to maximize Youden’s index (sensitivity + specificity − 1) for distinguishing PD from PR. To further characterize the discriminatory information contained in the pretreatment features, exploratory receiver operating characteristic (ROC) analyses were performed for lesion volume alone, and the combined volume–vascular field representation, and volume with territorial anatomy using a random forest model. Sensitivity, specificity, and AUC were reported to characterize lesion-level discrimination.

#### 2.5.3 Complete response

The association between pretreatment lesion volume and CR was modeled using univariable binary logistic regression with log-transformed pretreatment volume. For descriptive visualization, lesions were grouped into quantile-based bins according to log-transformed volume, and the observed CR proportion within each bin was displayed with 95% confidence intervals obtained by patient-level bootstrap resampling. The additional discriminatory information provided by vascular-field and arterial-territory variables beyond pretreatment lesion volume was evaluated using patient-grouped cross-validated ROC analysis, with performance summarized by the area under the ROC curve.

## 3 Results

### 3.1 Study cohort and early response distribution

The two source datasets represented clinically distinct populations of patients with brain metastases. The published UCSF-BMSR cohort included 306 patients undergoing Gamma Knife radiosurgery, with a mean age of 61 ± 12 years; 177 (57.8%) were women. Lung cancer was the most common primary malignancy (41.7%), followed by breast cancer (23.3%) and melanoma (12.6%). The published MOLAB cohort included 75 patients from five institutions: the mean age at the first recorded MRI was 56.8 ± 10.7 years, and 47 (62.7%) were women. Primary malignancies were predominantly lung cancer (43/75, 57.3%), followed by breast cancer (22/75, 29.3%) and melanoma (6/75, 8.0%). After application of the longitudinal imaging and lesion-level eligibility criteria, the present analysis included 116 patients with 513 brain metastases, comprising 85 patients with 471 lesions from UCSF and 31 patients with 42 lesions from MOLAB (Table 1).

**Table 1.**
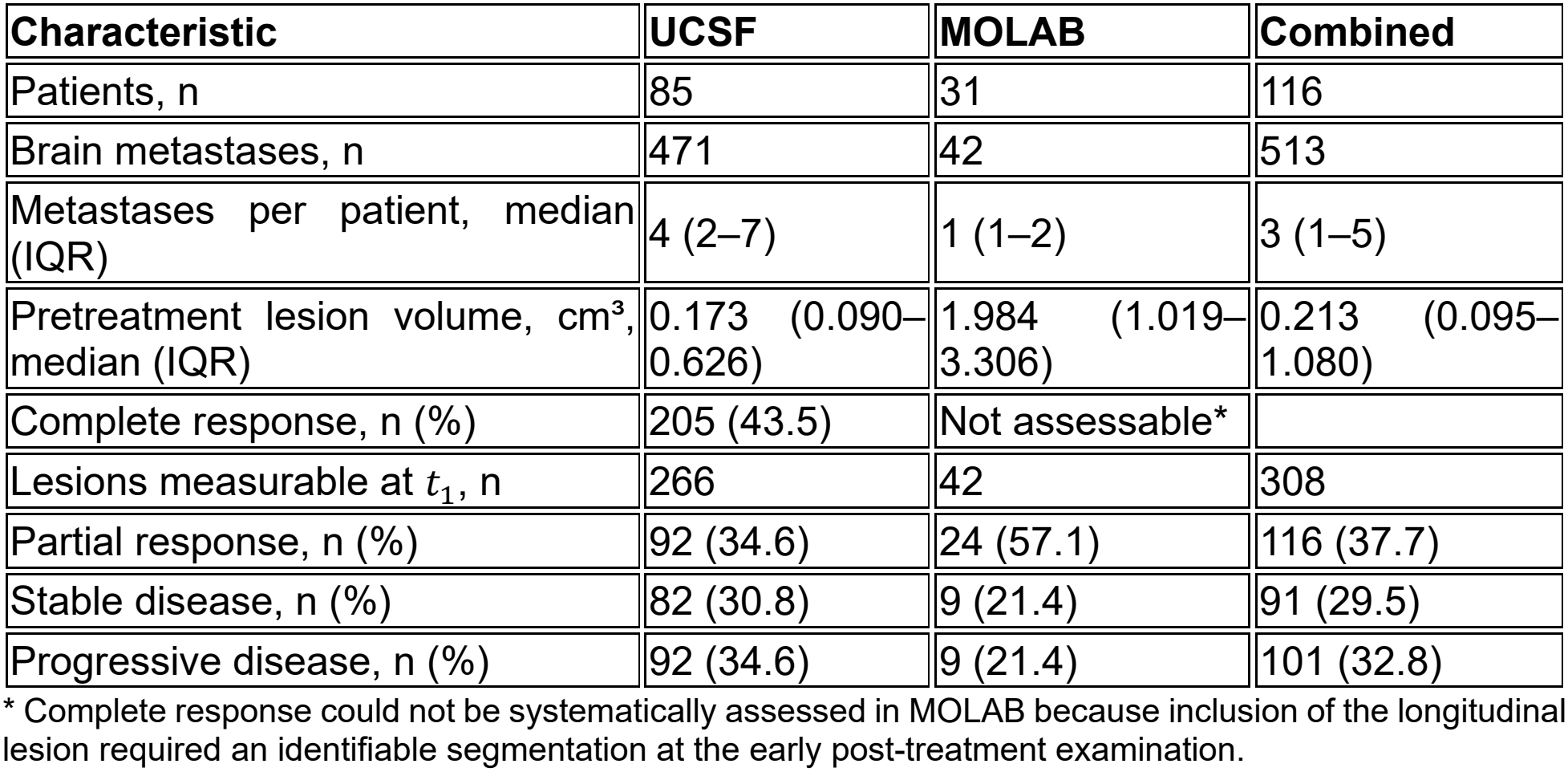
Lesion-level characteristics and early post-SRS response distribution in the UCSF and MOLAB longitudinal cohorts.

An important difference between the cohorts was lesion selection. The MOLAB SRS cohort was restricted to lesions meeting conventional RANO-BM measurability criteria, whereas the UCSF cohort represented a broader spectrum of metastases treated with Gamma Knife SRS, including many subcentimeter lesions. Among the 471 UCSF lesions, only 200 (42.5%) met the conventional RANO-BM measurable-lesion size criterion. Of the 266 UCSF lesions that remained radiographically identifiable at the early post-treatment examination, application of our volumetric response definitions yielded a relatively balanced distribution of PR (92/266, 34.6%), SD (82/266, 30.8%), and PD (92/266, 34.6%) (Table 1). In contrast, when RANO-BM–style linear response thresholds were applied to these same lesions, 181 (68.0%) were classified as SD, compared with 32 (12.0%) as PR and 53 (19.9%) as PD.

### 3.2 Peritumoral vascular field differs between volume-matched PD and PR lesions

To examine vascular spatial context independently of baseline lesion burden, 51 volume-matched PD–PR lesion pairs were compared. The paired analysis showed that the vascular association was strongest in the immediate peritumoral region, with the clearest separation observed within the 0–5 mm shell (Table 2). The association attenuated at greater distances from the lesion, indicating that the relevant vascular signal was spatially localized rather than broadly regional.

**Table 2.** Volume-matched comparison of peritumoral vascular fields between PD and PR lesions across distance-weighting kernels and spatial scales.

| Analysis | Vascular-field definition | PD > PR, n/N (%) | Two-sided *P*-value |
|---|---|---|---|
| Distance weighting, 0–5 mm shell | 1/*r* | 31/51 (60.8) | 0.061 |
|  | 1/*r*<sup>2</sup> | 32/51 (62.7) | 0.035 |
|  | **1/*r*<sup>3</sup>** | **33/51 (64.7)** | **0.029** |
| Spatial scale, 1/*r*<sup>3</sup> weighting | **0–5 mm** | **33/51 (64.7)** | **0.029** |
|  | 5–10 mm | 27/51 (52.9) | 0.257 |
|  | 10–20 mm | 29/51 (56.9) | 0.363 |
|  | 20–30 mm | 31/51 (60.8) | 0.177 |

Within the 0–5 mm shell, stronger inverse-distance weighting preserved the PD–PR difference while increasingly emphasizing vascular probability in close proximity to the lesion. On this basis, the 0–5 mm,1/*r*^3^-weighted vascular field was carried forward as the primary vascular metric for subsequent lesion-level analyses.

### 3.3 Peritumoral vascular field is associated with the magnitude of early progression, but not partial response

The subset of patients with non-zero vascular field included 146 measurable lesions (123 UCSF and 23 MOLAB), comprising 53 PD, 56 PR, and 37 SD lesions.

Within this cohort, the relationship between the pretreatment 0–5 mm, 1/*r*^3^ -weighted vascular field and subsequent volumetric change differed according to response direction (Figure 2A). For lesions with PD, increasing vascular-field values were associated with progressively greater relative volume increase, with a significant positive linear trend (*P* = 0.0106). In contrast, no corresponding relationship was observed among lesions with PR (*P* = 0.813). The binned lesion-level distributions were consistent with this asymmetric pattern, indicating that local vascular context was associated with the magnitude of early progression but not with the degree of tumor shrinkage.

**Figure 2.**
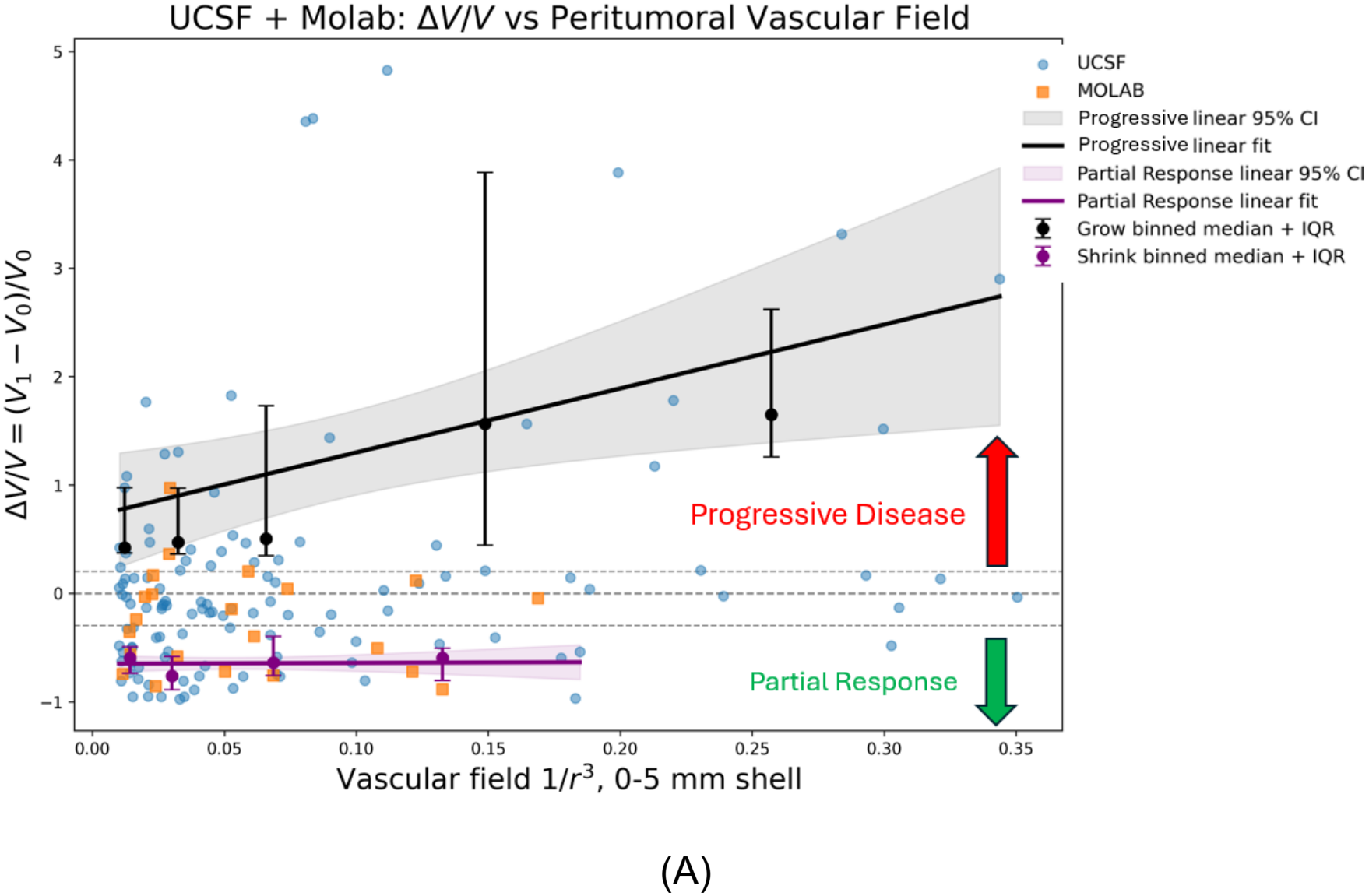
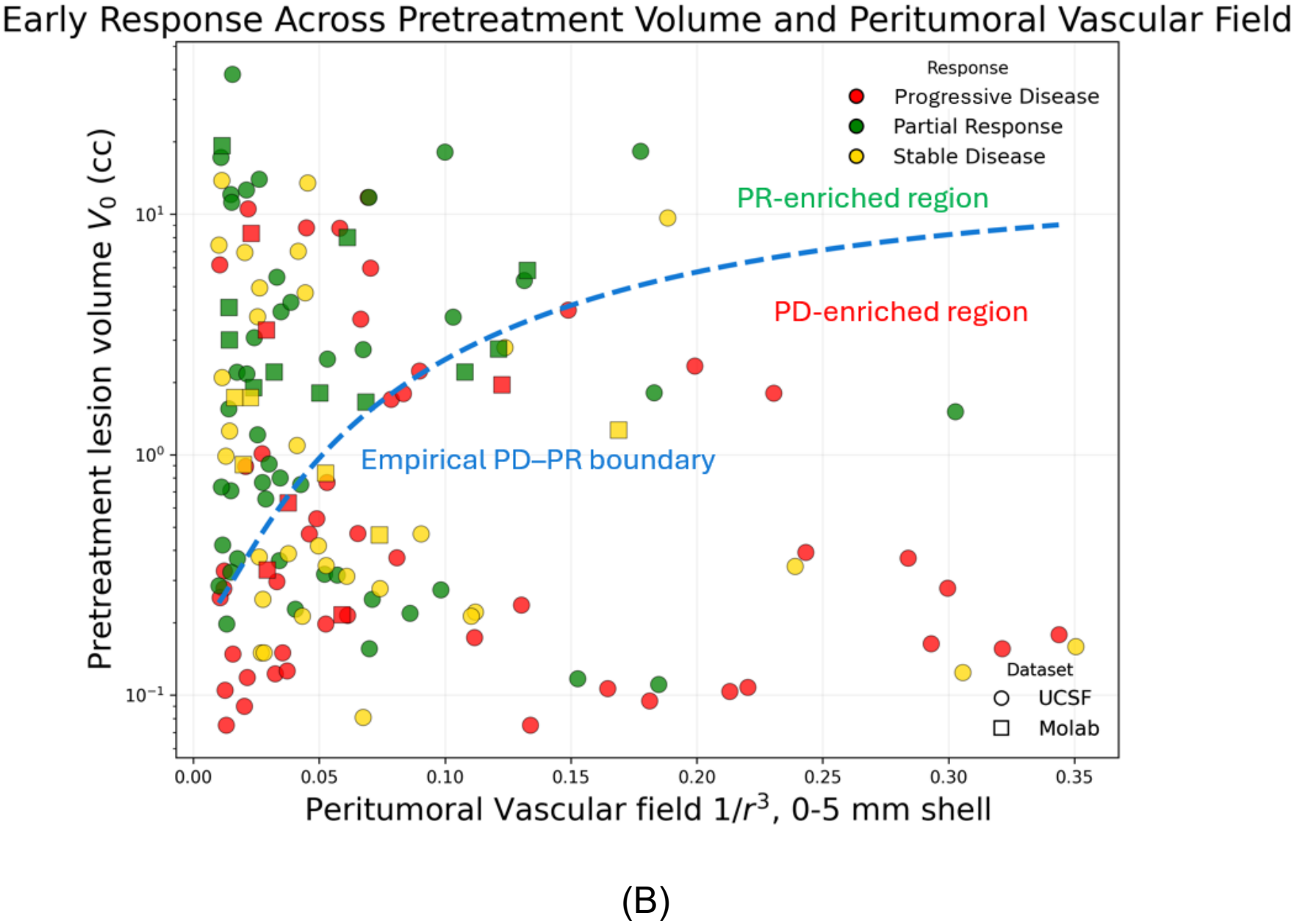
Pretreatment peritumoral vascular context is associated with early post-SRS lesion response. (A) Relative volumetric change from pretreatment to early follow-up (Δ*V*/*V*_0_) as a function of the pretreatment 0–5 mm, 1/*r*^3^ -weighted peritumoral vascular field in the UCSF and MOLAB cohorts. Among lesions with progressive disease (PD), increasing vascular-field values were associated with greater volumetric enlargement, whereas no corresponding relationship was observed among lesions with partial response (PR). Solid lines show linear fits with 95% confidence intervals. Binned medians and interquartile ranges are shown for descriptive visualization. Horizontal dashed lines indicate the prespecified volumetric response thresholds. (B) Joint distribution of pretreatment lesion volume and peritumoral vascular field according to early response category. The dashed blue curve represents an empirical PD–PR boundary and illustrates complementary organization of response across lesion burden and local vascular context, with PR-enriched and PD-enriched regions on opposite sides of the boundary. Marker shape denotes dataset and color denotes response category.

When pretreatment lesion volume and vascular field were considered jointly (Figure 2B), PR and PD lesions occupied partially distinct regions of the two-dimensional response space. An empirical PD–PR boundary separated a PR-enriched region from a PD-enriched region, illustrating that the relationship between response and vascular context depended on baseline lesion burden rather than on vascular field alone. Stable-disease lesions occupied an intermediate and overlapping distribution.

Because the empirical boundary was intended as a descriptive representation of the response landscape rather than a validated predictive classifier, its operating characteristics were summarized secondarily: Among the 109 PR and PD lesions included in the boundary analysis, the combined volume–vascular boundary showed higher sensitivity than the volume-only threshold: 0.77 (95% CI: 0.65-0.87) vs 0.64 (95% CI: 0.51-0.75) with *P* = 0.04 , while specificity was similar (0.73, 95% CI: 0.60-0.83 vs 0.73, 95% CI: 0.60-0.83). These findings support pretreatment lesion volume and local vascular context as complementary, rather than interchangeable, dimensions of early post-SRS response.

In lesion-level discrimination analysis, pretreatment volume alone achieved an AUC of 0.715 for distinguishing PD from PR (Figure 3). Incorporation of the peritumoral vascular field increased the AUC to 0.773 (ΔAUC = 0.058, 95% CI = 0.02-0.1, *P* < 0.01), whereas addition of territorial anatomy information did not improve discrimination (AUC, 0.587). These findings further support the complementary information provided by local vascular spatial context rather than territorial anatomy.

**Figure 3.**
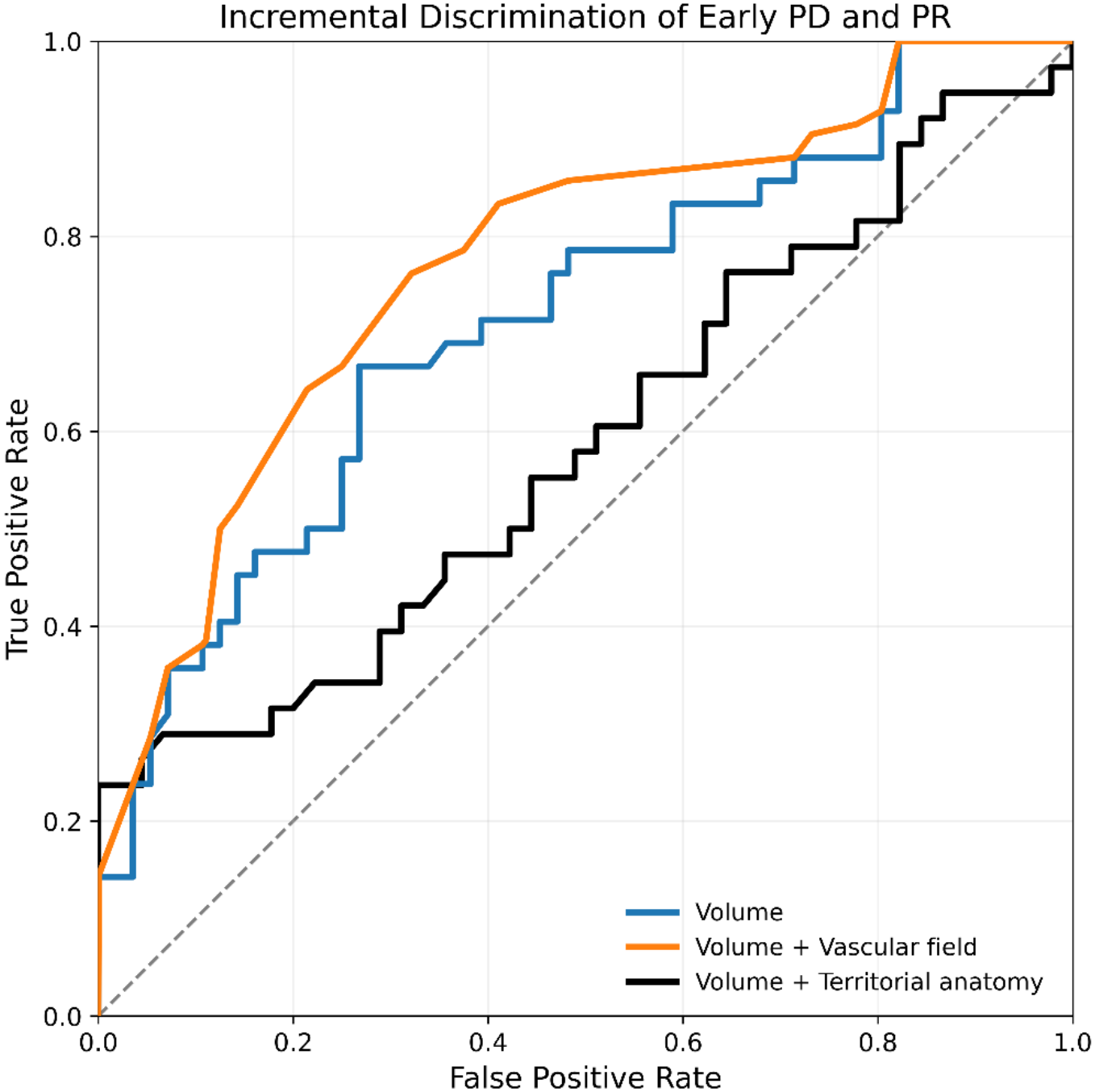
Exploratory lesion-level discrimination of early progressive disease and partial response using pretreatment lesion characteristics. Receiver operating characteristic curves are shown for pretreatment lesion volume alone, volume combined with the peritumoral vascular field, and volume combined with territorial anatomy information.

### 3.4 Complete response is predominantly associated with pretreatment lesion volume

The probability of CR decreased progressively with increasing pretreatment lesion volume (Figure 4A). A logistic model based on log-transformed pretreatment volume closely followed the observed binned CR rates, which declined from approximately two-thirds among the smallest lesions to less than 20% among larger lesions. This strong inverse relationship indicates that early complete radiographic response was primarily associated with baseline lesion burden.

**Figure 4.**
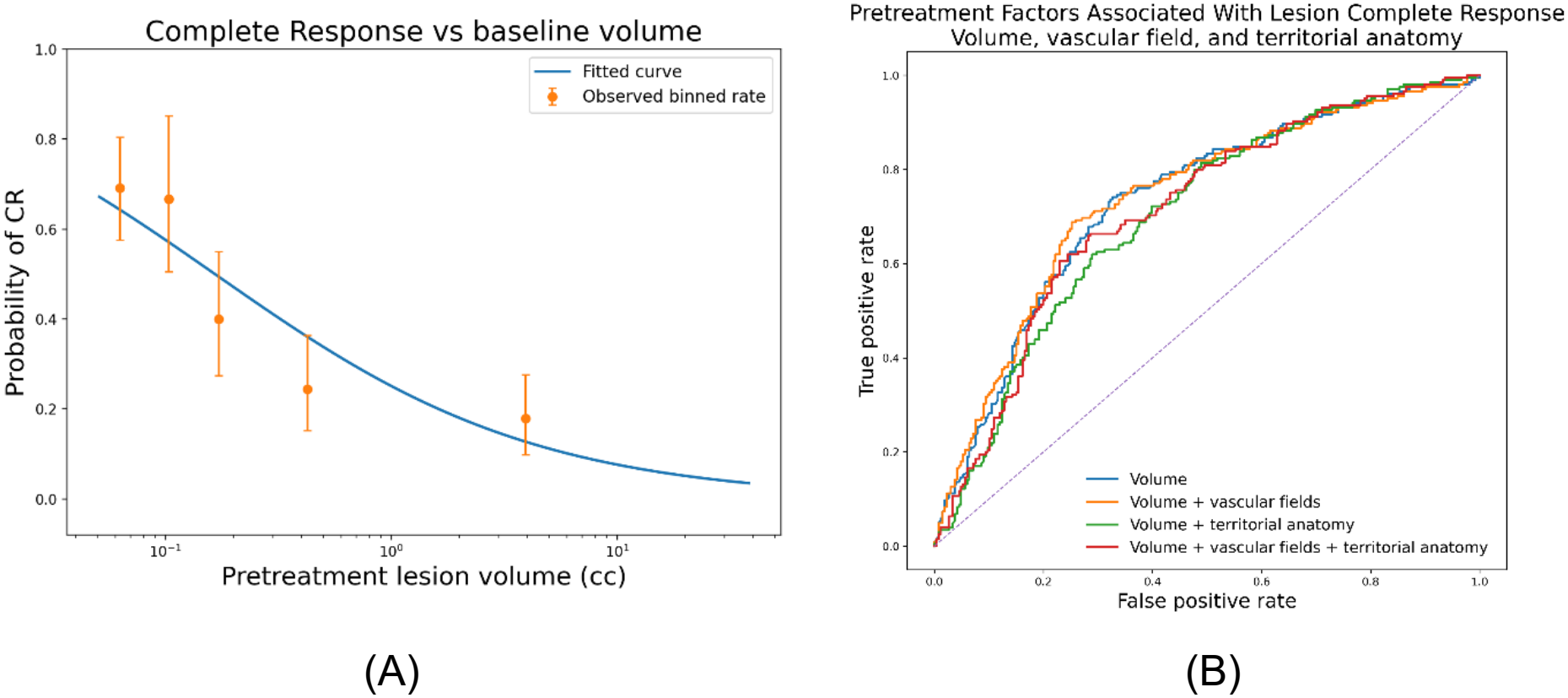
Pretreatment lesion volume is the dominant determinant of early complete response after stereotactic radiosurgery. (A) Observed complete response (CR) rates across pretreatment lesion-volume strata with 95% patient-level bootstrap confidence intervals. The fitted curve was obtained using univariable binary logistic regression with log-transformed pretreatment lesion volume. **(B)** Receiver operating characteristic curves evaluating discrimination of CR using pretreatment volume alone and after addition of vascular-field and territorial anatomy information. Cross-validation was grouped at the patient level to prevent lesions from the same patient from contributing to both training and validation folds. Addition of vascular-field or territorial anatomy information provided little improvement beyond pretreatment lesion volume alone.

Pretreatment lesion volume alone provided substantial discrimination for CR in patient-grouped cross-validation (AUC, 0.734; Figure 4B). Adding individual vascular-field measures, combined vascular-field measures, or territorial anatomy information produced no statistically significant improvement in discrimination, with all ROC curves remaining closely overlapping. These findings indicate that, unlike the response behavior of lesions remaining measurable after SRS, complete radiographic response was predominantly associated with pretreatment lesion volume rather than vascular spatial context.

## 4. Discussion

Our study contributes several elements to the study of brain metastases response after radiosurgery. The first is a large-scale demonstration of the increased discriminatory power of volumetric response assessment for tracking response assessment compared with standard RANO as has been previously reported in smaller cohorts[15]. Meanwhile, volumetric studies have similarly shown that three-dimensional assessment can evaluate more brain metastases, classify response more frequently, and identify progression earlier than conventional RANO-BM [15]. RANO-BM was developed primarily to standardize response assessment in clinical trials and defines measurable target lesions as at least 10 mm in longest diameter, with PR and PD determined by ≥30% decrease and ≥20% increase, respectively, in the sum of longest diameters[1, 12]. This framework is poorly aligned with the small metastases commonly encountered in modern SRS, as a substantial proportion of treated lesions fall below conventional measurability thresholds. Beyond lesion eligibility, unidimensional change is also suboptimal for characterizing early three-dimensional lesion evolution: for an approximately spherical lesion, a 20% increase in diameter corresponds to a 73% increase in volume, whereas a 30% decrease in diameter corresponds to a 66% reduction in volume. Consistent with this limitation, conventional linear response thresholds compressed much of the early post-SRS lesion evolution into the SD category. We therefore retained the clinically familiar CR/PR/SD/PD terminology while applying prespecified volumetric definitions to individual lesions and retaining continuous Δ*V*/*V*_0_ for trajectory analysis. These definitions should be regarded as an adaptation of RANO-BM rather than volumetric equivalents of its unidimensional criteria, allowing inclusion of small SRS-treated metastases while preserving sensitivity to early lesion-level change.

Within this lesion-level volumetric framework, the present study identifies a distinct relationship between pretreatment lesion burden, local cerebrovascular spatial context, and early response of brain metastases after SRS. As a primary novel contribution of our work, we demonstrate that among lesions that remained measurable after treatment, the relationship with vascular context was response-direction dependent: the magnitude of enlargement in PD increased with the pretreatment peritumoral vascular field, whereas the degree of shrinkage in PR was essentially independent of vascular field. This suggests a probabilistic mechanism of early PD – also often referred to as pseudo-progression-regulated by mechanisms involving arterial vasculature response to radiation. We also demonstrate that responses (both partial complete radiographic response) do not associate with arterial vessel field. As was previously known [16, 17], PR/CR was strongly associated with pretreatment lesion volume. Together, these findings suggest that baseline tumor burden and the surrounding vascular environment contribute to different components of early post-SRS response. This interpretation is consistent with Digernes et al.’s research identified pretreatment peritumoral vascular dysfunction associated with subsequent pseudo-progression using advanced perfusion and vessel architectural MRI, although the relevant peritumoral analysis was limited to 15 metastases [18].

The spatial localization of this association strengthens this interpretation. In the volume-matched PD–PR analysis, the clearest vascular difference was observed within the immediate 0–5 mm peritumoral region and weakened as progressively more distant tissue was considered. This finding argues against the vascular association simply reflecting a broadly vascular brain region. Instead, it points toward the local tumor–host interface as the relevant spatial scale. The 1/*r*^3^-weighted field was used to emphasize this immediate environment, but the biological interpretation does not depend on the mathematical kernel itself. Rather, the important observation is that vascular information closest to the lesion carried the strongest relationship with subsequent response. The limited contribution of territorial anatomy provides complementary evidence: knowing the broader arterial region in which a metastasis resided was less informative than characterizing its continuous spatial relationship to the local cerebrovascular architecture.

Several methodological considerations should be acknowledged. Firstly, cerebrovascular context was derived from a population-based atlas rather than patient-specific vascular imaging. Although patient-specific vascular characterization may provide greater anatomical precision, it would generally require dedicated vascular imaging sequences that are not routinely acquired in standard SRS workflows. Secondly, detailed lesion-level dose distributions were not consistently available across both cohorts. Dose may contribute to post-SRS response, although for the small metastases commonly treated with SRS, prescription and target coverage are typically relatively standardized. Incorporation of lesion-level dosimetry in future studies will nevertheless be important to determine whether treatment-related factors further modify the observed relationship between vascular context and response.

A more important clinical consideration is that early radiographic PD in this study should not be equated with pathologically confirmed viable tumor progression. During the first several months following SRS, lesion enlargement may reflect persistent or recurrent tumor, pseudo-progression, radiation-related vascular injury, necrosis, edema, or mixtures of these processes. The present study was intentionally designed to characterize this early volumetric phenotype rather than to adjudicate its underlying mechanism. Accordingly, the observed association between vascular field and enlargement identifies the pretreatment vascular environment as a potential contributor to the biological processes underlying early post-SRS enlargement. A related limitation is the absence of sufficiently long longitudinal follow-up to determine the subsequent course of these lesions. We could not establish whether vascular-associated enlargement later resolved, remained clinically manageable, or ultimately required salvage intervention. Thus, the present study identifies a pretreatment phenotype associated with early enlargement, but does not yet define the full longitudinal trajectory or clinical consequence of that phenotype.

This limitation directly defines the next stage of investigation. Future studies should link pretreatment volume–vascular characteristics with longer-term lesion trajectories and, most importantly, the subsequent need for salvage intervention. At present, the clinical significance of an enlarging post-SRS lesion often becomes apparent only through serial follow-up: brain MRI is generally obtained every 2–3 months after treatment, while pseudo-progression may occur from approximately 6 weeks to 15 months after SRS and radiation necrosis commonly emerges 6–24 months after treatment.[1, 19]. Conventional MRI cannot reliably distinguish treatment-related change from true progression, and advanced imaging approaches such as perfusion MRI, spectroscopy, and amino-acid PET may improve diagnostic confidence but do not provide a universally definitive solution [20, 21]. Thus, distinguishing a transient imaging change from a lesion that will ultimately require salvage treatment may require many months of observation, even with the use of advanced imaging. If the present pretreatment volume–vascular phenotype can be linked prospectively to these longer-term clinical outcomes, it could provide lesion-level risk stratification before SRS rather than after the response trajectory has already declared itself. Such information could support risk-adapted surveillance, with higher-risk lesions monitored more closely, and ultimately provide a basis for lesion-specific treatment planning or treatment adaptation.

## 5. Conclusion

Pretreatment lesion volume and local cerebrovascular spatial context appear to characterize different components of early response after SRS for brain metastases. Complete radiographic response was primarily associated with baseline lesion volume, while the magnitude of partial response showed little dependence on vascular context. In contrast, among lesions with early progressive enlargement, greater pretreatment peritumoral vascular field was associated with greater subsequent volume increase, with the strongest relationship localized to the immediate peritumoral region. When considered jointly, lesion volume and vascular field defined complementary partial-response–enriched and progression-enriched states before treatment. These findings support a lesion-level tumor–host framework for understanding early post-SRS response heterogeneity and provide a foundation for future studies linking pretreatment vascular context to long-term lesion outcome, salvage intervention, and ultimately individualized surveillance and treatment strategies.

## Data Availability

All data produced in the present study are available upon reasonable request to the authors

https://imagingdatasets.ucsf.edu/dataset/1

https://molab.es/datasets-brain-metastasis-1/?type=metasrd

